# Lack of evidence for genetic association of saposins A, B, C and D with Parkinson’s disease

**DOI:** 10.1101/2020.05.31.20061010

**Authors:** Yuri Ludwig Sosero, Sara Bandres-Ciga, Sharon Hassin-Baer, Roy N. Alcalay, Ziv Gan-Or, on behalf of the International Parkinson’s Disease Genomics Consortium (IPDGC)

## Abstract

The *PSAP* gene encodes prosaposin, which is later cleaved into four active saposins: saposin A, B, C and D. Mutations in these enzymes have been linked to specific lysosomal storage disorders. Recently, a genetic association between mutations in saposin D and Parkinson’s disease (PD) has been reported. To further examine whether variants in saposin D or the other saposins could be associated with Parkinson’s disease, we performed Optimized Sequence Kernel Association Test (SKAT-O) in 4,132 Parkinson’s disease patients and 4,470 controls. Furthermore, we analyzed data from a PD Genome Wide Association Study (GWAS) to examine the association of common variants in the *PSAP* locus with Parkinson’s disease risk (analysis on 56,308 patients) and age at onset (analysis on 28,568 patients). We did not find any statistically significant associations between neither rare nor common variants in saposin D, nor any of the other saposins, and PD risk or onset. These results suggest that PSAP variants play either a very minor role, or more likely, no role, in PD.

---

Sir,

We read with much interest the paper by Oji *et al.*, 2020, which suggested an association between mutations in the saposin D region of *PSAP* and Parkinson’s disease. In their paper, the authors have shown that variants in saposin D only partially segregate with Parkinson’s disease in three families of Japanese origin. For example, in family 1, at least three carriers of the *PSAP* p.Q453P variant did not have Parkinson’s disease, including one of the parents who was an obligate carrier. The authors also report an association of six common intronic variants within the saposin D region of the gene when comparing Japanese Parkinson’s disease patients and East Asian controls. Among these six variants, rs4747203 and rs885828 remained significant also when comparing the combined Japanese-Taiwanese Parkinson’s disease cohorts with East-Asian controls. These two variants are in perfect linkage disequilibrium (LD, r^2^=1) in East-Asians and represent the same allele. The authors further demonstrated, using different models, that *PSAP* mutations may affect prosaposin localization in the lysosome, autophagy, progressive motor decline in mice, and that patient-derived fibroblasts and dopaminergic neurons had α-synuclein accumulation.

*PSAP* encodes prosaposin, which is later cleaved into four active saposins: Saposin A, B, C and D. Saposins A-D are lysosomal hydrolases required for the degradation of sphingolipids (Kishimoto *et al*., 1992; Lieberman *et al*., 2011), and mutations in saposins may lead to accumulation of these sphingolipids. Saposins A, B, C, and in some case reports, saposin D (Diaz-Font *et al*., 2005; Devi *et al*., 2019), have been associated with specific lysosomal storage disorders (Kishimoto *et al*., 1992). In particular, saposin C activates glucocerebrosidase, a lysosomal enzyme encoded by *GBA*. Biallelic mutations in *GBA* and in rare cases in *PSAP* cause Gaucher disease, a lysosomal storage disorder. *GBA* mutations are also present in up to 20% of patients with Parkinson’s disease. However, a previous study suggested that *PSAP* variants are not associated with Parkinson’s disease. In this study it was shown that the specific saposin regions A-D, except for a nominal association of the saposin C region (with two variants identified, rs747170456 and rs745951723), were not associated with Parkinson’s disease (Bencheikh *et al*., 2018).

To further examine whether variants in saposin D or the other saposins are associated with Parkinson’s disease, we analyzed large cohorts of Parkinson’s disease patients and controls, (details on the specific cohorts are in Supplementary Table 1). These cohorts included a total of 4,132 Parkinson’s disease patients and 4,470 controls with full sequencing data on *PSAP*, and were used for analyzing the burden of rare variants in *PSAP* and saposins A-D separately. We further analyzed data on *PSAP* variants from the recent Parkinson’s disease genome-wide association study (GWAS) on 37,688 cases, 18,618 UK Biobank proxy-cases (i.e., subjects with a first degree relative with Parkinson’s disease) and 1.4 million controls, all Europeans (Nalls *et al*., 2019). To examine whether variants in the *PSAP* locus are associated with age at onset (AAO) of Parkinson’s disease, we analyzed data from a recent GWAS on the AAO of Parkinson’s disease, with data on 28,568 patients (Blauwendraat *et al*., 2019). The study protocol was approved by the institutional review board at McGill University.

The cohorts analyzed at the NIH (Supplementary Table 1) were captured using TruSeq DNA or TruSeq DNA PCRFree sample preparation kits, followed by whole genome sequencing (WGS). The cohorts analyzed at McGill University were captured using Molecular Inversion Probes (MIPs), which specifically target the regions of interest, as we previously described (Ross *et al*., 2016). A complete pipeline for MIPs design and a supplementary table including the MIPs used for the sequencing are available in Bencheikh *et al*., 2018, and are also available upon request. In both NIH and McGill cohorts, variants were filtered for minor allele frequency < 0.01, genotype missingness (< 0.05 in NIH cohorts; <0.10 in McGill cohorts) and Hardy–Weinberg equilibrium (P <= 1E-6). In addition, in the NIH cohorts, quality control for mean sequence depth (< 25x) and contamination rate (> 3%) of the samples was performed. In the McGill cohorts, separate analyses with different depth of coverage thresholds (15x, 30x and 50x) were performed. Our study has power > 80% to detect statistically significant (*p* < 0.05) rare variants frequency difference of 1% in one group vs. 2% in the second group (patients or controls), and power of 100% to identify the effects of rs4747203 and rs885828 with *p* < 0.05 as reported by Oji *et al*. We tested the association of rare variants in saposin D and the other saposins with Parkinson’s disease using optimized sequence Kernel association tests (SKAT-O). These burden tests were first done on all rare variants, followed by analyses only with variants that are potentially functional (nonsynonymous, splice-site, frameshift and stop variants).

In the analyses of both McGill (with 15x, 30x and 50x thresholds) and NIH cohorts, SKAT-O results showed no association between *PSAP* variants and Parkinson’s disease. Furthermore, when we separately analyzed saposins A-D, no association was found (Table 1). Of note, no known pathogenic saposin D mutations were found in either McGill or NIH cohorts. To examine the association of common variants in the *PSAP* locus with Parkinson’s disease in individuals of European origin, and specifically the association of rs4747203 and rs885828, we extracted data from the most recent GWASs on Parkinson’s disease risk and age at onset (Blauwendraat *et al*., 2019; Nalls *et al*., 2019). No *PSAP* variants passed the significance threshold for an association with risk or AAO of Parkinson’s disease (Figure 1). In particular, the two SNPs identified by Oji *et al*. as significant both in the Japanese and in the combined Taiwanese-Japanese cohorts, rs4747203 and rs885828 (Oji *et al*., 2020), showed lack of association with risk of Parkinson’s disease (p = 0.7577, OR = 1.0032, 95% CI = 1.0239-0.9828 and *p* = 0.7288, OR = 0.996, 95% CI = 1.017-0.976, respectively) or with its AAO (p = 0.0164, b = 0.2367, SE = 0.0986 and *p* = 0.01679, b = −0.2358, SE = 0.0986, respectively. These *p* values are not corrected for multiple comparisons and are not statistically significant after Bonferroni correction). Of note, these two SNPs are in perfect Linkage Disequilibrium (r^2^=1, D=1) both in East-Asian and European populations, implicating that they represent the same allele (https://ldlink.nci.nih.gov). Since Oji *et al*. compared Japanese Parkinson’s disease patient population to East-Asian controls, which include multiple ethnicities, we further compared the frequencies of these two variants with Japanese controls from a previously published GWAS (Satake *et al*., 2009, data also found in Oji *et al*., 2020 Supplementary table 8). The alternative allele frequency of rs885828 (which is in perfect LD with rs4747203) was 0.7 in healthy Japanese controls, very similar to the frequency in Japanese Parkinson’s disease patients reported by Oji *et al*. study, 0.72 (*p* = 0.1, χ^2^ = 2.732).

**Table 1.** Association of rare saposin variants with Parkinson’s disease

|  | **P-value** |  |  |  |
| --- | --- | --- | --- | --- |
|  | **SKAT-O**<br>**McGill All** | **SKAT-O**<br>**McGill Func** | **SKAT-O NIH**<br>**All** | **SKAT-O NIH**<br>**Func** |
| **Sap A** | 0.40 | 0.39 | 0.34 | NA |
| **Sap B** | 0.13 | 0.15 | 0.79 | NA |
| **Sap C** | 0.76 | 0.76 | NA | NA |
| **Sap D** | 0.72 | 0.42 | 0.47 | NA |
| ***PSAP*** | 0.19 | 0.16 | 0.42 | 0.67 |

SKAT-O, Optimized Sequential Kernel Association Test; Func, functional variants, including nonsynonymous, frame-shift, splice site and stop variants; Sap, saposin; *PSAP*, prosaposin; NA, not applicable, not enough variants for analysis.

**Figure 1.**
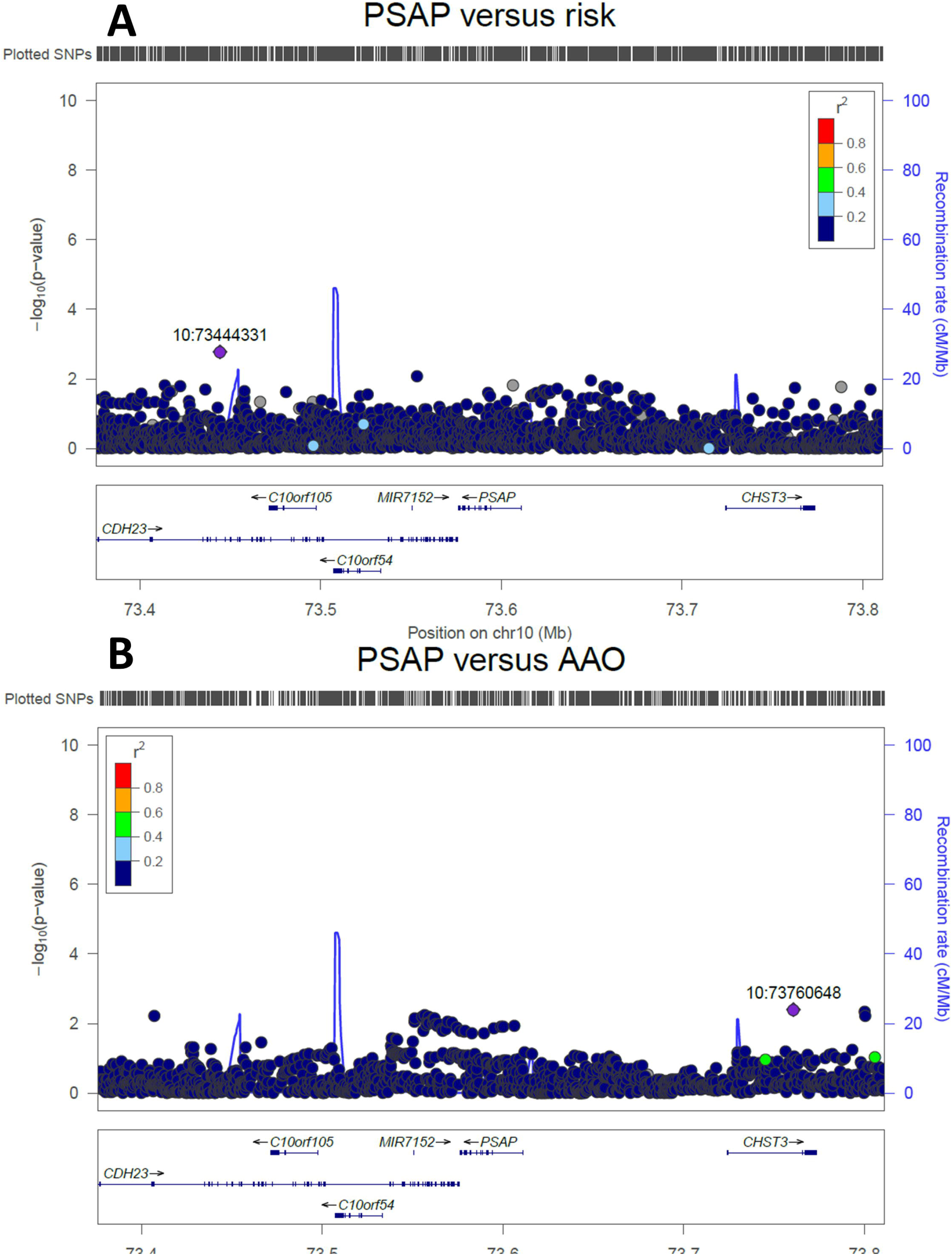
Zoom plots of common genetic variants in *PSAP* and their association with Parkinson’s disease. **A)** Zoom plot of variants within *PSAP* and their association with risk of Parkinson’s disease. Data are taken from the recent Parkinson’s disease genome-wide association study (GWAS), which included 37,688 cases, 18,618 UK Biobank proxy-cases and 1.4 million controls (Nalls *et al*., 2019). None of the variants in *PSAP* were associated with risk of Parkinson’s disease. **B)** Zoom plot of variants within *PSAP* and their association with age at onset (AAO) of Parkinson’s disease. Data are from the recent GWAS on the AAO of Parkinson’s disease, which included 28,568 patients (Blauwendraat *et al*., 2019). None of the variants in *PSAP* were associated with AAO of Parkinson’s disease.

In conclusion, our findings do not support an association between either rare or common variants in saposin D or any of the other saposins, and Parkinson’s disease. We cannot completely rule out the possibility of very rare pathogenic saposin D variants associated Parkinson’s disease, although we did not find an association between functional variants and PD. It is possible that differences in the populations tested could have led to the differences between our findings and those reported by Oji *et al*. However, the current results suggest that *PSAP* variants play either a very minor role, or more likely, no role at all in Parkinson’s disease.

## Data Availability

All data is available upon reasonable request from the corresponding author.

## Acknowledgements

We would like to thank participants for their contribution to the study. ZGO is supported by the Fonds de recherche du Québec - Santé (FRQS) Chercheurs-boursiers award, in collaboration with Parkinson Quebec, and by the Young Investigator Award by Parkinson Canada. We would like to thank Jennifer A. Ruskey, Farnaz Asayesh, Uladzislau Rudakou, Lynne Krohn, Eric Yu, Kheireddin Mufti, Helene Catoire and Vessela Zaharieva for their assistance. We would also like to thank all members of the International Parkinson Disease Genomics Consortium (IPDGC). For a complete overview of members, acknowledgements and funding, please see the Supplemental material and/or http://pdgenetics.org/partners. We would like to thank the Accelerating Medicines Partnership initiative (AMP-PD) for the publicly available whole genome sequencing data including cohorts from the Michael J. Fox Foundation (MJFF) and National Institutes of Neurological Disorders and Stroke (NINDS) BioFIND study, Harvard Biomarkers Study (HBS), the NINDS Parkinson’s disease Biomarkers Program (PDBP), and MJFF Parkinson’s Progression Marker Initiative (PPMI). Finally, we would like to thank the 23andMe research participants who contributed to this work and the 23andMe Research Team: Michelle Agee, Stella Aslibekyan, Adam Auton, Robert K. Bell, Katarzyna Bryc, Sarah K. Clark, Sarah L. Elson, Kipper Fletez-Brant, Pierre Fontanillas, Nicholas A. Furlotte, Pooja M. Gandhi, Karl Heilbron, Barry Hicks, David A. Hinds, Karen E. Huber, Ethan M. Jewett, Yunxuan Jiang, Aaron Kleinman, Keng-Han Lin, Nadia K. Litterman, Matthew H. McIntyre, Kimberly F. McManus, Joanna L. Mountain, Sahar V. Mozaffari, Priyanka Nandakumar, Elizabeth S. Noblin, Carrie A.M. Northover, Jared O’Connell, Aaron A. Petrakovitz, Steven J. Pitts, G. David Poznik, J. Fah Sathirapongsasuti, Janie F. Shelton, Suyash Shringarpure, Chao Tian, Joyce Y. Tung, Robert J. Tunney, Vladimir Vacic, Xin Wang.

## Supplementary material

Supplementary Table 1

